# PRKCA overexpression is frequent in young oral tongue squamous cell carcinoma patients and is associated with poor prognosis

**DOI:** 10.1101/2020.06.21.20122648

**Authors:** Thomas Parzefall, Julia Schnoell, Laura Monschein, Elisabeth Foki, David Tianxiang Liu, Alexandra Frohne, Stefan Grasl, Johannes Pammer, Trevor Lucas, Lorenz Kadletz, Markus Brunner

**Author notes:** Corresponding author at: Medical University of Vienna, Department of Otorhinolaryngology, Head and Neck Surgery, Waehringer Guertel 18-20, 1090 Vienna, Austria.

## Abstract

Oral tongue squamous cell carcinoma (OTSCC) have an increasing incidence in young patients and many have an aggressive course of disease. The molecular mechanisms for this increase are unknown and biologic markers to identify high risk patients are lacking.

In an unbiased data screening for differential protein expression of younger (≤45 years) and older (>45 years) OTSCC patients in The Cancer Genome Atlas (TCGA) cohort (n=98) we identified Protein kinase C alpha (PRKCA), to be significantly more frequently overexpressed in younger versus older patients (p=0.0001). These results were experimentally validated and confirmed in an independent Austrian OTSCC patient sample (n=34) by immunohistochemistry (p=0.0026). PRKCA upregulation was associated with negative anamnesis for alcohol consumption (p=0.009) and tobacco smoking (p=0.02). Univariate and multivariate analysis of overall survival (OS) and disease-free survival (DFS) showed a significantly worse prognosis in patients with tumors overexpressing PRKCA regarding OS (univariate p= 0.04, multivariate p< 0.01). In the young subgroup both OS and DFS were significantly decreased in PRKCA positive patients (both p< 0.001). TCGA messenger RNA enrichment analysis showed 24 mRNAs with significant differential expression in PRKCA positive OTSCC (all p≤ 0.05 after Benjamini-Hochberg correction).

Our findings suggest the potential existence of a distinct molecular subtype of alcohol and tobacco negative, high risk OTSCC in a significant proportion of early onset individuals. Our findings warrant validation in additional OTSCC patient cohorts. Further analysis of the molecular PRKCA interactome may decipher the underlying mechanisms of carcinogenesis and clinicopathological behavior of PRKCA overexpressing OTSCC.

## Introduction

Head and Neck squamous cell carcinoma (HNSCC) are the 6^th^ most common cancer, with a worldwide annual incidence of more than 800,000 cases ^1^. Well-established risk factors for the development of HNSCC are chronic alcohol and tobacco consumption, leading to mutagenesis, chromosomal instability and progressive epithelial dysplasia and carcinoma formation ^2–4^. HNSCC typically have a peak incidence around the 6^th^ to 7^th^ decade of life ^5,6^.

While the incidence of tobacco and alcohol related HNSCC decreased over the last decades in many western countries, human papilloma virus (HPV)-associated carcinomas of the oropharynx have significantly increased ^7^ and are typically diagnosed around the age of 50.

A notable worldwide increase is also observed in oral tongue squamous cell carcinoma (OTSCC) among young individuals ^8–10^. Interestingly, this increase can neither be explained by an increased tobacco and alcohol exposure, nor by virus-driven tumorigenesis, suggesting additional pathomechanisms in this particular patient subgroup ^6,11–13^.

While the data on prognosis and biological behavior of OTSCC in young patients in comparison to older age groups is still ambiguous, younger patients have been reported to have higher rates of regional and distant metastasis and a highly aggressive course of disease in recurrent cases ^14–16^.

The identification of specific molecular markers in early onset OTSCC patients may help to clarify the mechanism of disease and clinicopathologic tumor behavior in this age group, identify high risk patients and allow improvement in treatment strategies.

The increasing availability of high throughput DNA and RNA sequencing technology in recent years has led to a number of comparative studies that have investigated genomic and transcriptomic aberrations in young and older OTSCC samples. The overall mutational spectrum so far has shown no remarkable differences between young and older patients ^17,18^. However, one study reported an association of high DNA copy number variant (CNV) burden with a reduced overall survival within the subgroup of young patients analyzed ^19^. Additionally, young OTSCC patients have been found to differ in mRNA expression pattern of immunomodulatory markers such as LAG3 and HAVCR2 when compared to older patients ^20^.

On the tissue level, numerous proteins have been evaluated as molecular prognostic markers using immunohistochemistry or targeted tissue micro-arrays in OTSCC samples. A focus has been set on previously known cell cycle, angiogenesis and epithelial-mesenchymal transition related proteins ^21–26^. However, the reported results are variable and do not allow strong conclusions and studies focusing on young OTSCC patients are scarce.

In contrast to targeted investigations of pre-selected proteins or protein sets, large-scale proteomics approaches using mass spectrometry allow a broad and unbiased examination of tumor protein expression and detection of novel protein markers. The Cancer Genome Atlas (TCGA) database (https://www.cancer.gov/tcga) provides a unique data set that includes clinical and proteomics data from different cancer tissue samples, including OTSCC.

The objective of this study was to identify candidate molecular markers associated with early onset OTSCC and poor prognosis. For this purpose, we used TCGA data to screen for protein markers differentially expressed in young OTSCC patients compared to older patients. The data gained from this initial exploratory TCGA screening served as a base for further targeted evaluation of protein expression in a local Austrian OTSCC patient sample and upstream genomic and transcriptomic *in silico* analysis.

## Patients and Methods

### Study design and setting

The study was designed as a two-step retrospective observational cohort study. In the first step, protein markers associated with young age (≤ 45 years) in patients with OTSCC were explored within the TCGA databank. In the second step, identified candidate protein markers during step one were experimentally validated in an independent OTSCC cohort treated at an Austrian tertiary referral center.

The main outcome measure was the statistical association of any candidate protein marker with patient age equal or below 45 years. The secondary outcome measures were the statistical association of any identified marker with alcohol and tobacco smoking anamnesis and the overall and disease-free survival.

### TCGA data retrieval and selection

TCGA Head and Neck Squamous Cell carcinoma data sets were retrieved via the cBio cancer portal ^27^. Samples from all OTSCC patients with proteomic (RPPA and z-scores), mRNA, DNA and clinical data available were selected for analyses. Data were extracted from three distinct datasets within TCGA (Head and Neck Squamous Cell Carcinoma, TCGA, Provisional; Head and Neck Squamous Cell Carcinoma TCGA PanCancer Atlas; Head and Neck Squamous Cell Carcinoma, TCGA, Nature 2015). In the case of overlapping patients in the data sets, duplicate samples were excluded from the analysis. Data were initially sorted by patient age at onset of disease and divided into 2 groups of young (≤45 years) and older (>45 years) patients for further analysis in accordance with previous studies related to early onset OTSCC ^9,14,20^.

### Austrian patient sample

Tissue samples from newly diagnosed, previously untreated patients with oral tongue squamous cell carcinoma (OTSCC) obtained during primary surgical resection or diagnostic panendoscopy at the Department of Otorhinolaryngology, Head and Neck Surgery, Medical University of Vienna, between 1999 and 2016 were retrieved from the local tissue archive. All samples with an availability of a sufficient tissue volume to obtain 3 slides of 4 micrometers (1x primary antibody immunostaining, 1x isotype control, 1x hematoxylin/eosin only) were included in the analysis. Patients with a previous history of malignant disease or an additional concurrent primary malignancy were excluded.

Additionally, clinical parameters including age, sex, AJCC tumor staging (7^th^ edition), treatment modalities, history of alcohol and/or tobacco consumption and clinical outcome were extracted from the patient records.

The study was conducted in concordance with the WMA Declaration of Helsinki and was approved by the Ethics Committee of the Medical University of Vienna (approval no. 1262/2019).

### Smoking and alcohol consumption status in the patient samples

For statistical analysis, tobacco smoking and alcohol consumption status was coded in a binary fashion.

Patients with a self-reported lifelong cumulative smoking history of less than 100 cigarettes were coded as non-smokers, in concordance with the NHI National Cancer Institute definitions (https://cdebrowser.nci.nih.gov/cdebrowserClient/cdeBrowser.html#/search?publicId=2181650&version=1.0). Current and reformed smokers with a cumulative dose exceeding 100 cigarettes were considered smokers.

Regarding alcohol status, subjects with a consumption of less or equal to 2 alcoholic drinks/week (social and never-drinkers) were considered alcohol-negative. Three or more alcoholic drinks per week were considered a positive alcohol anamnesis.

Patients with unknown status in tobacco and alcohol consumption were not included in the analysis.

### Immunohistochemistry

Immunohistochemical (IHC) staining of archived formalin fixed and paraffin embedded tissue sections was performed using a Lab Vision Ultra kit (Thermo Fisher Scientific, Waltham, MA, USA) according to the manufacturer’s protocol. Initially, the ideal antibody dilutions (1:800, Rabbit MAB Anti-PRKCA, AB no. ab32376, Abcam, Cambridge, UK and 1:800, Mouse MAB Anti-ANXA1, AB no. EH17a, Developmental Studies Hybridoma Bank, University of Iowa, IA, US) and retrieval buffer (citrate buffer) were assessed using human cerebral and esophageal samples, respectively. These samples were also used as positive controls. Tissue samples were dewaxed and rehydrated using xylol, ethanol and water. Endogenous peroxidase activity was blocked in three percent H_2_O_2_ for 15 minutes. Antigen retrieval was performed in a microwave (600 W) using citrate buffer (pH 6.0). Subsequently, Ultra V Block was applied for 5 minutes. Then the tissue samples were incubated with the primary antibodies against PRKCA and ANXA1 at room temperature for one hour. Next, the primary antibody enhancer and horseradish peroxidase enhancer were applied for 10 and 15 minutes, respectively. Antibody staining was visualized using the UltraVision Plus Detection System DAB Plus Substrate System (Thermo Fisher Scientific, Waltham, MA, USA) and samples were counterstained using hematoxylin Gill II (Merck, Darmstadt, Germany). For negative controls, primary antibody was replaced by rabbit immunoglobulin G isotype control (Abcam, Cambridge, UK).

### Protein expression quantification

In the TCGA cohort, protein expression fold changes with a z-score of ≥ +1.96 or ≤ −1.96 (p≤ 0.05) were considered as overexpression or underexpression, respectively.

In the Viennese OTSCC samples, semiquantitative analysis of immunohistochemically stained tissue sections was performed by two experienced pathologists (L.M., J.P.) who were blinded to the clinical patient data. Both, the fraction of positively stained carcinoma cells and the expression intensity was measured to classify protein expression levels. Samples were graded according to fraction of positive cells into 0 (<5% of cells positive), 1 (5-33% of cells positive), 2 (>33-66% of cells positive), 3 (>66% of cells positive) and according to staining intensity into 0 (none), 1 (weak), 2 (moderate), 3 (strong). Positive cell fraction score and intensity score were summed up to give a final IHC score of 0 (min.) to 6 (max.). A total IHC score of ≤3 was defined as low expression whereas a score ≥4 was considered overexpression.

For inter-group comparison, protein expression z-scores were calculated for the Viennese samples using the protein expression values, mean and standard deviation. As in the TCGA cohort, protein expression fold changes with a z-score of ≥ +1.96 or ≤ −1.96 (p≤ 0.05) were considered as upregulation or downregulation, respectively.

### Statistical analysis

2-tailed fisher’s exact test was used to determine the statistical association of target protein differential expression and age equal or below 45 years (statistical cut-off: p≤ 0.01) as well as alcohol/tobacco consumption behavior (positive versus negative consumption history) in both the TCGA cohort and the Vienna patient samples.

For survival analysis Kaplan Meier estimates were computed. Intergroup differences were assessed with log-rank tests.

Multivariate survival analysis on the combined cohort (TCGA+ Vienna) was performed using a cox-regression model including patient age, T-classification, N-classification and PRKCA protein expression status.

All statistical calculations were carried out with Stata (StataCorp, College Station, TX, USA) and Prism GraphPad (GraphPad Software, San Diego, CA, USA).

Enrichment analysis of mRNA expression and comparative genomic analysis between PRKCA positive and PRKCA negative TCGA samples was conducted with the integrated sample comparison function of the cBio cancer portal ^27^ with the student’s t-test. Correction for multiple testing was performed with the Benjamini-Hochberg procedure with an accepted false detection rate of 5%. Therefore, a q-value ≤ 0.05 was considered a significant association.

## Results

### Patient characteristics

Within the TCGA HNSCC data sets a total of 98 OTSCC samples contained information about both clinical parameters and proteomics data and were included in the analysis (Supplemental Table 1).

The patient characteristics of the TCGA cohort and the Viennese sample are summarized in table 1. The TCGA cohort consisted of 63 male and 35 female patients ranging from 19 to 87 years (mean: 57.5; SD: 13.6). 15 patients were 45 years or younger and 82 patients were older than 45 years. Age was unknown in 1 TCGA sample (TCGA-CQ-A4CA-01) which therefore was excluded in any age-related statistical calculation.

**Table 1:**
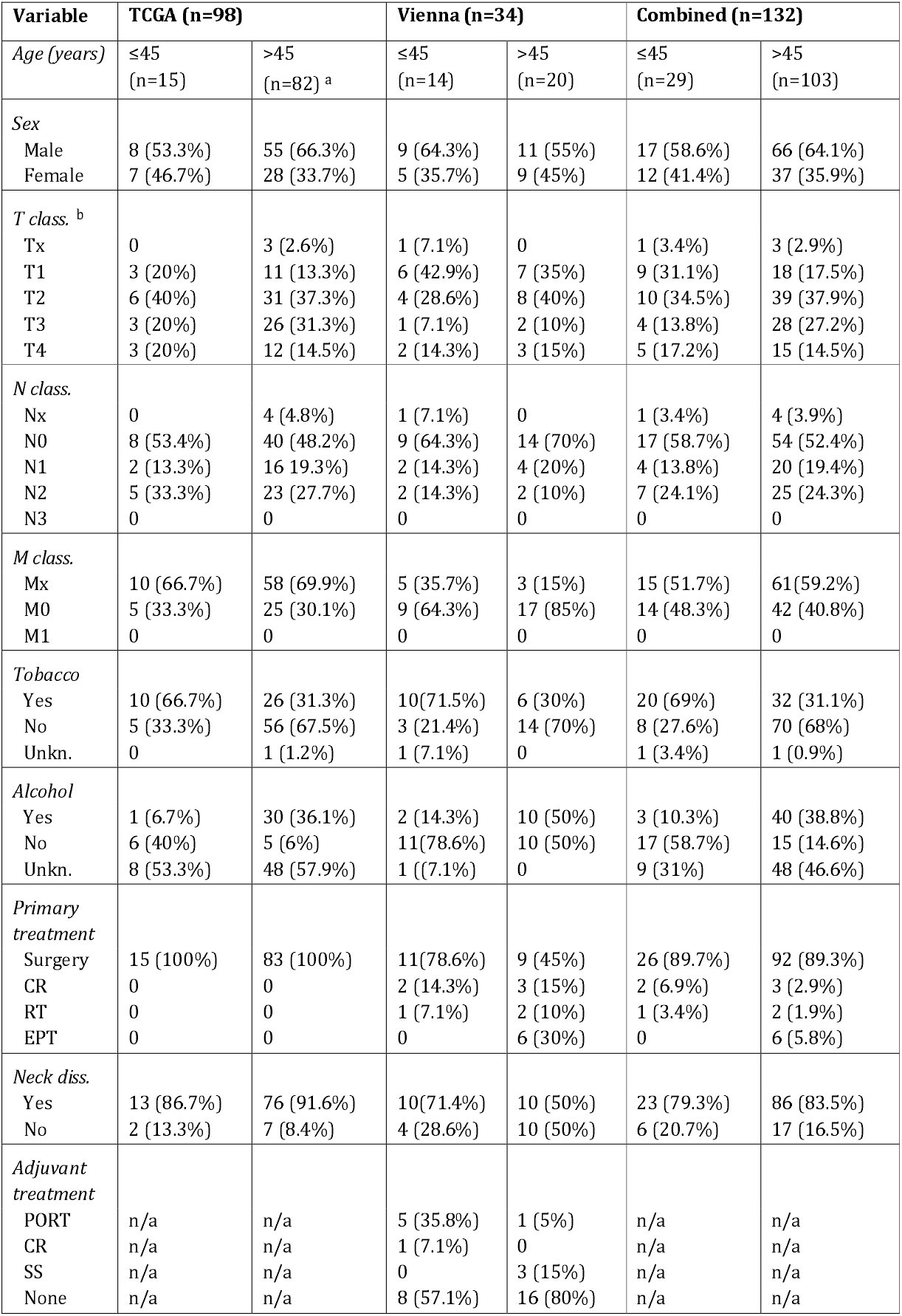
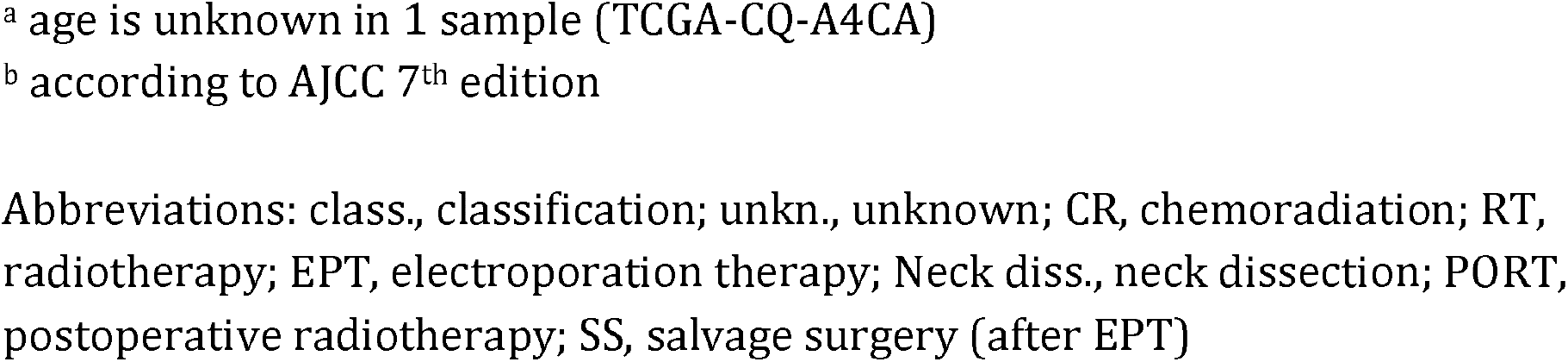
Clinicopathologic features of the study samples

| Variable | TCGA (n=98) |  | Vienna (n=34) |  | Combined (n=132) |  |
| --- | --- | --- | --- | --- | --- | --- |
| *Age (years)* | ≤45<br>(n=15) | >45<br>(n=82) <sup>a</sup> | ≤45<br>(n=14) | >45<br>(n=20) | ≤45<br>(n=29) | >45<br>(n=103) |
| *Sex* |  |  |  |  |  |  |
| Male | 8 (53.3%) | 55 (66.3%) | 9 (64.3%) | 11 (55%) | 17 (58.6%) | 66 (64.1%) |
| Female | 7 (46.7%) | 28 (33.7%) | 5 (35.7%) | 9 (45%) | 12 (41.4%) | 37 (35.9%) |
| *T class. <sup>b</sup>* |  |  |  |  |  |  |
| Tx | 0 | 3 (2.6%) | 1 (7.1%) | 0 | 1 (3.4%) | 3 (2.9%) |
| T1 | 3 (20%) | 11 (13.3%) | 6 (42.9%) | 7 (35%) | 9 (31.1%) | 18 (17.5%) |
| T2 | 6 (40%) | 31 (37.3%) | 4 (28.6%) | 8 (40%) | 10 (34.5%) | 39 (37.9%) |
| T3 | 3 (20%) | 26 (31.3%) | 1 (7.1%) | 2 (10%) | 4 (13.8%) | 28 (27.2%) |
| T4 | 3 (20%) | 12 (14.5%) | 2 (14.3%) | 3 (15%) | 5 (17.2%) | 15 (14.5%) |
| *N class.* |  |  |  |  |  |  |
| Nx | 0 | 4 (4.8%) | 1 (7.1%) | 0 | 1 (3.4%) | 4 (3.9%) |
| N0 | 8 (53.4%) | 40 (48.2%) | 9 (64.3%) | 14 (70%) | 17 (58.7%) | 54 (52.4%) |
| N1 | 2 (13.3%) | 16 (19.3%) | 2 (14.3%) | 4 (20%) | 4 (13.8%) | 20 (19.4%) |
| N2 | 5 (33.3%) | 23 (27.7%) | 2 (14.3%) | 2 (10%) | 7 (24.1%) | 25 (24.3%) |
| N3 | 0 | 0 | 0 | 0 | 0 | 0 |
| *M class.* |  |  |  |  |  |  |
| Mx | 10 (66.7%) | 58 (69.9%) | 5 (35.7%) | 3 (15%) | 15 (51.7%) | 61 (59.2%) |
| M0 | 5 (33.3%) | 25 (30.1%) | 9 (64.3%) | 17 (85%) | 14 (48.3%) | 42 (40.8%) |
| M1 | 0 | 0 | 0 | 0 | 0 | 0 |
| *Tobacco* |  |  |  |  |  |  |
| Yes | 10 (66.7%) | 26 (31.3%) | 10 (71.5%) | 6 (30%) | 20 (69%) | 32 (31.1%) |
| No | 5 (33.3%) | 56 (67.5%) | 3 (21.4%) | 14 (70%) | 8 (27.6%) | 70 (68%) |
| Unkn. | 0 | 1 (1.2%) | 1 (7.1%) | 0 | 1 (3.4%) | 1 (0.9%) |
| *Alcohol* |  |  |  |  |  |  |
| Yes | 1 (6.7%) | 30 (36.1%) | 2 (14.3%) | 10 (50%) | 3 (10.3%) | 40 (38.8%) |
| No | 6 (40%) | 5 (6%) | 11 (78.6%) | 10 (50%) | 17 (58.7%) | 15 (14.6%) |
| Unkn. | 8 (53.3%) | 48 (57.9%) | 1 (7.1%) | 0 | 9 (31%) | 48 (46.6%) |
| *Primary treatment* |  |  |  |  |  |  |
| Surgery | 15 (100%) | 83 (100%) | 11 (78.6%) | 9 (45%) | 26 (89.7%) | 92 (89.3%) |
| CR | 0 | 0 | 2 (14.3%) | 3 (15%) | 2 (6.9%) | 3 (2.9%) |
| RT | 0 | 0 | 1 (7.1%) | 2 (10%) | 1 (3.4%) | 2 (1.9%) |
| EPT | 0 | 0 | 0 | 6 (30%) | 0 | 6 (5.8%) |
| *Neck diss.* |  |  |  |  |  |  |
| Yes | 13 (86.7%) | 76 (91.6%) | 10 (71.4%) | 10 (50%) | 23 (79.3%) | 86 (83.5%) |
| No | 2 (13.3%) | 7 (8.4%) | 4 (28.6%) | 10 (50%) | 6 (20.7%) | 17 (16.5%) |
| *Adjuvant treatment* |  |  |  |  |  |  |
| PORT | n/a | n/a | 5 (35.8%) | 1 (5%) | n/a | n/a |
| CR | n/a | n/a | 1 (7.1%) | 0 | n/a | n/a |
| SS | n/a | n/a | 0 | 3 (15%) | n/a | n/a |
| None | n/a | n/a | 8 (57.1%) | 16 (80%) | n/a | n/a |

<sup>a</sup> age is unknown in 1 sample (TCGA-CQ-A4CA)
<sup>b</sup> according to AJCC 7<sup>th</sup> edition
Abbreviations: class., classification; unkn., unknown; CR, chemoradiation; RT, radiotherapy; EPT, electroporation therapy; Neck diss., neck dissection; PORT, postoperative radiotherapy; SS, salvage surgery (after EPT)

The Viennese sample consisted of 34 (20 male, 14 female) patients with an age range of 20 to 75 years (mean: 49.5; SD: 15.9) at the time of first diagnosis. 14 patients were 45 years or younger and 20 patients were older than 45 years.

### PRKCA is frequently overexpressed in young OTSCC patients

In the initial TCGA screening, two proteins, Protein kinase C alpha (PRKCA) and Annexin 1 (ANXA1) met the criteria for further experimental validation (Fisher’s exact two-tailed p≤ 0.01). Subsequently, ANXA1 overexpression was not statistically overrepresented in younger compared to older patients in the Viennese validation sample (p= 1.0). However, PRKCA was found to be significantly more frequently overexpressed in young (≤45years) compared to older (>45 years) patients in both the TCGA cohort (n= 97; p=0.0001) and the Vienna validation study sample (n=34; p= 0.002). (Figure 1)

**Figure 1:**
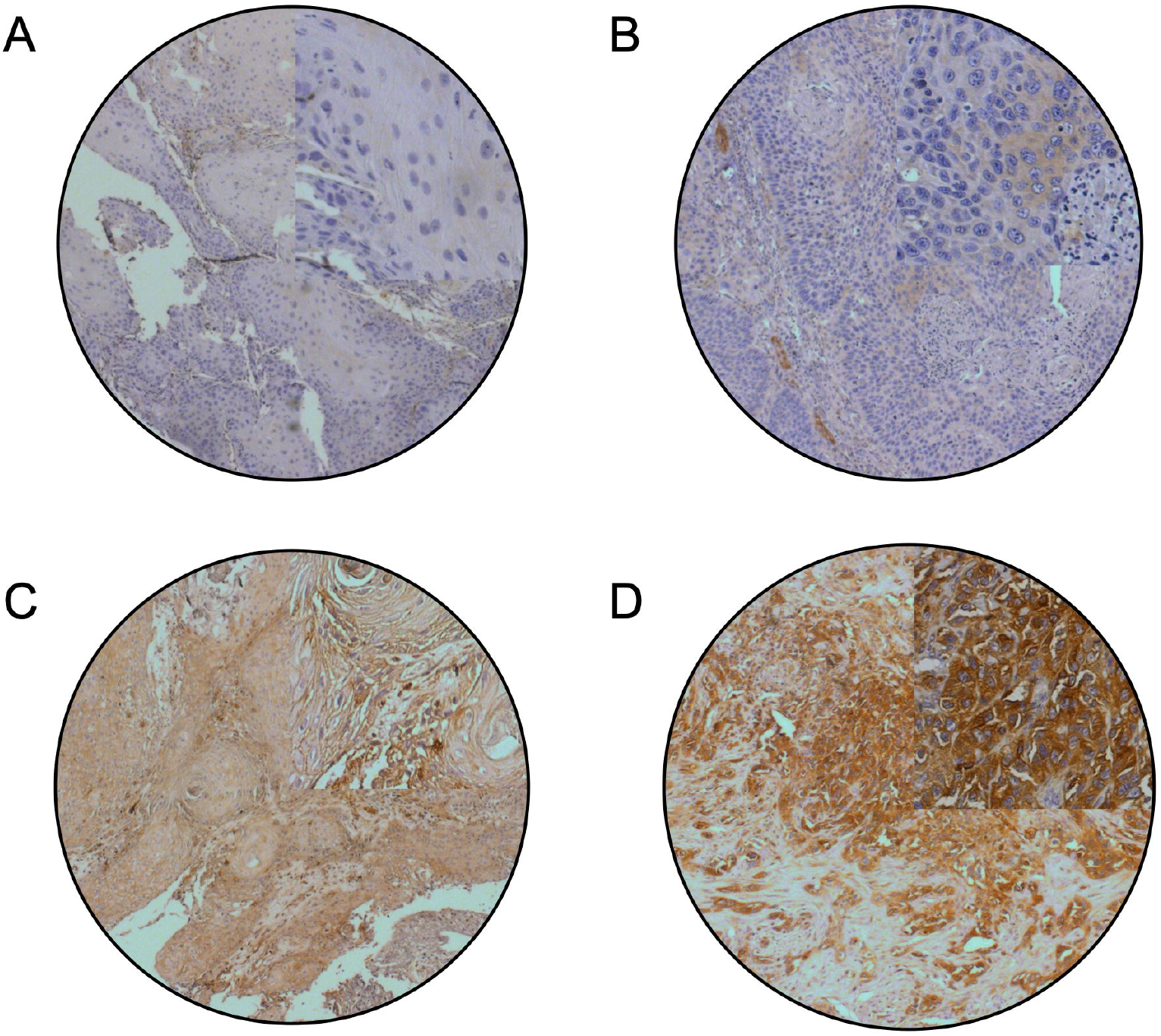
Representative immunohistochemistry images of OTSCC tissue stained for PRKCA (10x). Examples of OTSCC with A) no, B) weak focal, C) moderate and D) strong PRKCA immunoreactivity. Areas in the upper right quadrants were enlarged to 40x.

In the TCGA cohort, 6 out of 15 patients (40%) aged ≤45 years had a significant PRKCA overexpression with a z-score above +1.96, whereas among the patients >45 years, 2 out of 82 patients (2.4%) showed PRKCA upregulation.

Median PRKCA immunohistochemistry score in the Vienna patients ≤45 years was 0.5 (range 0-6). In the group > 45 years, the median total IHC score was 0 (range 0-3). Four out of 14 young patients (28.6%) compared to none of the patients > 45 years (0%) had a total IHC score of 4 or above and were defined as PRKCA upregulated.

### PRKCA overexpression is associated with adverse clinical outcome

To investigate whether differential expression of the candidate proteins PRKCA and ANXA1 had an influence on the oncologic outcome, we calculated the Kaplan Meier survival functions in the TCGA and Viennese sample sets (Figure 2 and Supplemental Figure 1). ANXA1 did not show an association with either OS or DFS in the total study population or within any subgroup (Supplemental Figure 1) and was therefore not considered further as a candidate prognostic marker.

However, PRKCA overexpression significantly correlated with adverse clinical outcome. In the Vienna sample, reduced overall (OS) and disease-free survival (DFS) and tumor recurrence at last follow-up were significantly associated with PRKCA upregulation in the total sample and the young patient fraction (all univariate p≤ 0.002). Similarly, in the TCGA cohort, DFS was significantly worse in young OTSCC patients overexpressing PRKCA (univariate p= 0.02). Combining the two study samples resulted in a significant association of PRKCA overexpression with poor DFS (univariate p<0.0001) and OS (univariate p< 0.001) in the young patients.

**Figure 2:**
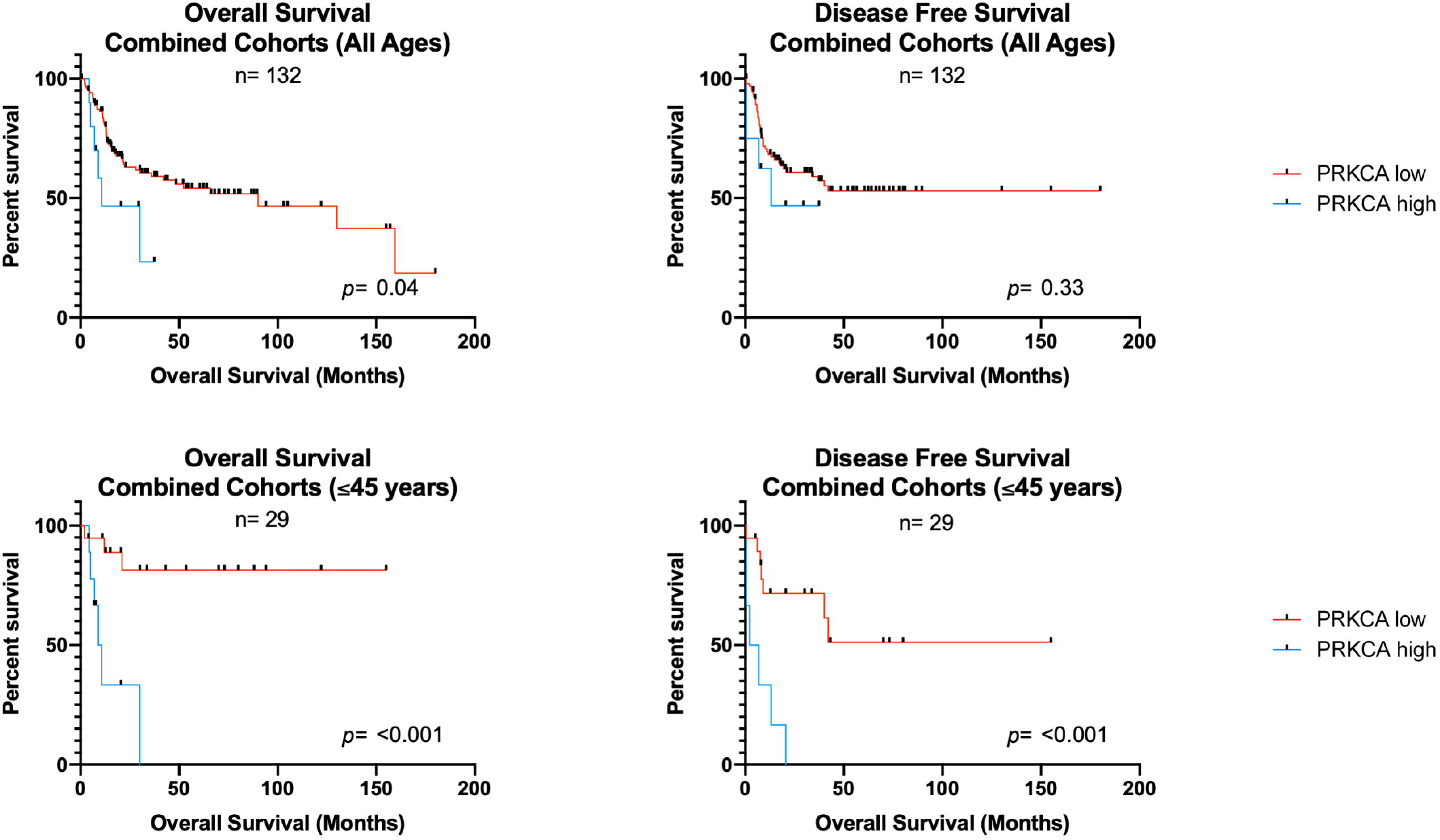
Kaplan Meier survival curves in correlation to PRKCA expression level. Univariate survival analysis shows significantly reduced OS of OTSCC patients with high PRKCA expression (p=0.04). In the subgroup of patients aged 45 years or younger, high PRKCA expression compromises both, OS and DFS (both p< 0.001).

Additionally, we performed univariate survival analysis for patient age, AJCC T-classification and N-classification in the combined cohort which showed a significant association of T-classification and N-classification (both univariate p <0.01) with OS. No significant additional association was found with DFS.

In the multivariate analysis, T-classification (p <0.01), N-classification (p= 0.05) and PRKCA overexpression (p <0.01) remained significantly associated with OS. None of the analyzed parameters were associated with DFS after multivariate testing.

A detailed list of survival data and calculated univariate and multivariate hazard ratios in regard to PRKCA is displayed in Table 2.

**Table 2:** Median overall and disease-free survival and hazard ratios in the patient groups

| Cohort | Median Survival (months) |  | UV HR (95% CI) | UV p-value <sup>a</sup> | MV HR (95% CI) | MV p-value <sup>b</sup> |
| --- | --- | --- | --- | --- | --- | --- |
|  | *PRKCA positive* | *PRKCA negative* |  |  |  |  |
| Vienna All OS | 15 | 130 | 34.34<br>(4.15 - 284.1) | 0.001 | - | - |
| Vienna Young OS | 15 | Undef. | 141.8<br>(12.24 - 1642) | <0.001 | - | - |
| Vienna All DFS | 0.5 | Undef. | 107.7<br>(10.03 - 1158) | <0.001 | - | - |
| Vienna Young DFS | 0.5 | Undef. | 31.15<br>(3.56 - 272.7) | 0.002 | - | - |
| Vienna All TU at LFU | 15 | Undef. | 53.07<br>(5.82 - 483.9) | <0.001 | - | - |
| Vienna Young TU at LFU | 15 | 94 | 44.25<br>(4.87 - 401.9) | <0.001 | - | - |
| TCGA DFS All | Undef. | Undef. | 0.58<br>(0.14 - 2.47) | 0.46 | - | - |
| TCGA OS All | Undef. | 52.27 | 1.51<br>(0.38 - 5.95) | 0.56 | - | - |
| TCGA OS Young | 10.74 | Undef. | 3.2<br>(0.47 - 21.6) | 0.23 | - | - |
| TCGA DFS Young | 6.83 | Undef. | 16.5<br>(1.69 - 161.2) | 0.02 | - | - |
| Combined OS All | 10.74 | 90.05 | 3.62<br>(1.06 - 12.36) | 0.04 | 3.6<br>(1.46-8.85) | < 0.01 |
| Combined DFS All | 13 | Undef. | 1.9<br>(1.06 - 12.36) | 0.33 | 1.71<br>(0.21-13.82) | 0.61 |
| Combined OS Young | 9.87 | Undef. | 19<br>(3.61 - 99.57) | <0.001 | - | - |
| Combined DFS Young | 4.5 | Undef. | 18.23<br>(3.56 - 93.30) | <0.001 | - | - |

<sup>a</sup> determined by log rank test
<sup>b</sup> determined by cox regression analysis
Abbreviations: PRKCA, protein kinase C alpha; UV, univariate; MV, multivariate; HR, hazard ratio; CI, confidence interval; OS, overall survival; DFS, disease free survival; TU, tumor; LFU, last follow-up; undef., undefined

### PRKCA overexpression is frequent in alcohol and tobacco negative OTSCC

To assess if PRKCA is differentially expressed in patient samples with or without alcohol and tobacco exposure, we calculated the statistical associations of PRKCA overexpression with both alcohol and tobacco consumption behavior of the patients. We found an association of PRKCA upregulation with a negative history of alcohol and tobacco consumption in both the Viennese sample and the TCGA cohort. In the TCGA cohort, this association was statistically significant for alcohol consumption (p= 0.01, 2-tailed). When combining the data from both the Vienna and the TCGA samples, the correlation between both alcohol consumption (p=0.009, 2-tailed) and tobacco smoking (p=0.02, 2-tailed) became statistically significant.

### Messenger RNA expression profiles differ significantly between PRKCA positive and PRKCA negative OTSCC

To identify potential upstream molecular alterations associated with PRKCA protein overexpression, we compared genomic and transcriptomic data of PRKCA positive and PRKCA negative TCGA samples. Curated mRNA expression and DNA sequence and copy number data were available from all except 1 OTSCC sample (TCGA-CQ-6221) in the HNSCC TCGA PanCancer Atlas dataset (n=97).

Comparative genomic analysis between PRKCA positive (n=8) and negative (n=89) samples within the TCGA database did not render a significant difference in the total mutational count or any specific DNA sequence or copy number variation pattern distinctive of either subgroup (data not shown).

However, mRNA enrichment analysis identified 24 mRNAs with a significant differential up-or down-regulation in the 2 subgroups. A significant overexpression was detected for 18 and underexpression for 6 mRNAs in the PRKCA positive samples (Table 3). Dysregulation of 13 of these differentially expressed mRNAs (TGM3, EPHB6, SCNN1A, TMPRSS11D, GRHL3, CLDN4, ATF4, RAB25, DHX32, VPS4B, GHITM, SLC25A25, DTX3L) is known to be associated with different human cancer types.

**Table 3:** Differentially expressed mRNA in PRKCA high (+) and PRKCA low (-) OTSCC samples in the TCGA dataset

| Gene | Mean log2 expression in PRKCA+ (SD) | Mean log2 expression in PRKCA- (SD) | Log Ratio | p-Value <sup>a</sup> | q-Value <sup>b</sup> | Higher expression in |
| --- | --- | --- | --- | --- | --- | --- |
| TPRG1 | 8.65 (0.21) | 7.04 (1.31) | 1.61 | 3.17e-14 | 4.46e-10 | PRKCA+ |
| CLDN4 | 12.63 (0.42) | 10.83 (1.71) | 1.80 | 3.07e-8 | 2.157e-4 | PRKCA+ |
| RAB25 | 12.07 (0.30) | 10.92 (1.40) | 1.15 | 4.08e-7 | 1.914e-3 | PRKCA+ |
| SCNN1A | 11.21 (0.52) | 9.41 (1.51) | 1.80 | 2.138e-6 | 7.521e-3 | PRKCA+ |
| DHX32 | 10.42 (0.12) | 10.04 (0.43) | 0.38 | 3.397e-6 | 8.391e-3 | PRKCA+ |
| ASPG | 7.61 (0.62) | 5.61 (2.02) | 2.00 | 4.424e-6 | 8.391e-3 | PRKCA+ |
| CYP2C18 | 9.38 (0.77) | 6.88 (2.19) | 2.50 | 5.060e-6 | 8.391e-3 | PRKCA+ |
| STIMATE | 7.01 (0.13) | 7.42 (0.45) | -0.42 | 5.087e-6 | 8.391e-3 | PRKCA- |
| TMPRSS11D | 12.14 (0.86) | 9.36 (2.60) | 2.78 | 5.368e-6 | 8.391e-3 | PRKCA+ |
| MANSC1 | 9.11 (0.34) | 8.04 (1.24) | 1.07 | 6.472e-6 | 9.106e-3 | PRKCA+ |
| PRRC2A | 12.08 (0.19) | 12.69 (0.45) | -0.61 | 1.049e-5 | 0.0134 | PRKCA- |
| DTX3L | 11.06 (0.22) | 11.70 (0.69) | -0.64 | 1.735e-5 | 0.0192 | PRKCA- |
| GRHL3 | 11.04 (0.50) | 9.60 (1.78) | 1.44 | 1.772e-5 | 0.0192 | PRKCA+ |
| TMEM246 | 8.78 (0.50) | 7.36 (1.92) | 1.42 | 2.446e-5 | 0.0245 | PRKCA+ |
| NCKAP5L | 8.56 (0.28) | 9.39 (0.64) | -0.84 | 2.614e-5 | 0.0245 | PRKCA- |
| ATF4 | 12.78 (0.13) | 12.41 (0.53) | 0.37 | 2.994e-5 | 0.0263 | PRKCA+ |
| TGM3 | 13.39 (1.85) | 7.97 (4.12) | 5.43 | 3.240e-5 | 0.0268 | PRKCA+ |
| SLC25A25 | 10.32 (0.41) | 9.13 (0.90) | 1.19 | 4.061e-5 | 0.0309 | PRKCA+ |
| EPHB6 | 8.88 (0.44) | 7.69 (1.59) | 1.18 | 4.170e-5 | 0.0309 | PRKCA+ |
| VPS4B | 11.08 (0.16) | 10.65 (0.62) | 0.43 | 4.626e-5 | 0.0325 | PRKCA+ |
| GHITM | 12.46 (0.17) | 11.98 (0.39) | 0.48 | 5.583e-5 | 0.0374 | PRKCA+ |
| ZNF579 | 6.35 (0.30) | 7.14 (0.91) | -0.79 | 6.626e-5 | 0.0424 | PRKCA- |
| LEXM | 6.40 (1.16) | 3.14 (2.00) | 3.27 | 8.219e-5 | 0.0493 | PRKCA+ |
| CALM3 | 11.84 (0.14) | 12.19 (0.40) | -0.35 | 8.406e-5 | 0.0493 | PRKCA- |

<sup>a</sup> student's t-test
<sup>b</sup> corrected p-value after Benjamini-Hochberg false detection rate correction procedure
Gene list ranked according to q-value (statistical significance level)

## Discussion

Over the past decades, a decline in the use of tobacco and alcohol in most Western countries has been accompanied by a decreasing incidence of most HNSCCs ^7–10^. Running counter to this trend, a concurrent rise in cases of early-onset OTSCC has been observed. Since OTSCC were shown not to be appreciably linked to HPV, the cause for this increase is still obscure ^12,13^. A set of clinical differences, including age, severity and exposure to established risk factors, have prompted the notion that early-onset and late-onset OTSCC may represent distinct disease subtypes. A higher CNV burden and attenuated anti-tumor immune activity, reflected by lower cytolytic activity scores, fewer neoantigens and mRNA-level downregulation of immunomodulators such as HAVCR2 and LAG3, have been linked to early-onset OTSCC ^19,20^. So far, however, reliable biomarkers are lacking, preventing molecular subtype characterization, development of tailored therapies and reliable prognoses.

In the present study, protein kinase C-alpha (PRKCA) was identified as a protein marker that is significantly overexpressed in early-onset OTSCC patients (aged 45 years or younger at diagnosis).

While previous studies aiming to identify biomarkers have focused on known pre-selected protein markers from other cancer types, we chose an unbiased approach with an initial exploratory screening of TCGA high-throughput proteomics data. Such an approach allows the detection of novel markers but is associated with a higher risk of false positives. Therefore, we validated the results by immunohistochemical analysis in a second, local OTSCC sample with a rigid protocol, in which the analyzing pathologists were blind to the clinical data of the patients. The combination of the local Viennese sample and the larger TCGA dataset also allowed to upscale the total sample size and statistical power and to level out sample heterogeneity inherent to retrospective data.

The role of PRKCA as a tumorigenic marker is conceivable. Protein kinase C isoforms have been long identified as the intracellular receptors of phorbol esters that promote tumor formation during 2-stage chemical-induced carcinogenesis in mouse skin ^28,29^. Subsequently, PRKCA has been shown to intersect with the MAPK/ERK and PI3K/AKT pathways, which are frequently active in several cancer types, promoting tumor progression by suppressing apoptosis and inducing proliferation, migration, invasion and angiogenesis ^30–36^. In addition to a more frequent upregulation in younger patients in this study, PRKCA expression correlated with a negative tobacco smoking and alcohol consumption anamnesis and was associated with reduced OS and DFS. While age itself did not have an influence on the prognosis, PRKCA overexpression status showed a highly significant association with poor OS and DFS in the young OTSCC subgroup (Figure 2). Based on these observations, we hypothesized that, rather than representing one subtype, early-onset OTSCC may be subdivided into high-risk PRKCA-positive and lower-risk PRKCA-negative forms. To further investigate this idea, TCGA mRNA-level expression data was retrieved and tested for differential expression between PRKCA-positive and PRKCA-negative tumors. Twenty-four genes were found to be significantly differentially expressed, including 13 genes that have previously been linked to cancer progression. The majority (n=12) had higher expression in the PRKCA-positive group and included multiple pro-invasive genes, which is consistent with the more aggressive course seen in these patients. PRKCA activates PI3K/AKT signaling which, downstream, promotes epithelial-to-mesenchymal transition (EMT), a process crucial for invasion and metastasis that is characterized by the loss of E-Cadherin and upregulation of pro-invasive factors such as vimentin and N-Cadherin ^32,37^. Overexpression of SCNN1A, TMPRSS11D and CLDN4, which have been suggested as biomarkers for poor prognosis in ovarian and cervical cancer, as well as GRHL3 that has been shown to induce EMT and promote cell migration and invasion was observed ^38–42^. Interestingly, TGM3 and EPHB6 had a higher expression in the PRKCA-positive group and are considered suppressors of EMT ^43,44^. EPHB6, however, plays a dual role in cancer as it also promotes cell proliferation and is overactive in aggressive triple-negative breast cancer ^45^. Further PRKCA-positive upregulated genes that have been reported in cancer include frequently dysregulated ATF4 and RAB25 as well as VPS4B, GHITM and SLC25A25 ^46–50^. The sole gene with higher expression in the PRKCA-negative group and a known role in cancer was DTX3L, which affects E-Cadherin expression over the FAK/PI3K/AKT pathway ^51^.

Based on our findings and the known molecular roles of PRKCA in relevant cancer pathways we propose a model for PRKCA overexpression as a driver in the tumorigenesis of a subset of OTSCC patients (Figure 3). However, the tumor initiator(s) remain elusive. Given the DNA mutational landscape does not differ significantly between PRKCA positive and negative tumors a specific genetic event or genetic predisposition, which is often found in young patients with malignant disease, seems unlikely. The rising incidence of early onset patients in recent decades would rather suggest a role for environmental, dietary or lifestyle factors, though alcohol, tobacco smoking and HPV do not play a substantial role. Interestingly, an emerging excess is also observed for lung adenocarcinoma (LADC) in recent decades in both the US and China, particularly among younger females and PRKCA overexpression is also associated with lower survival rates in LADC patients. ^52–54^ This trend is counter to a general reduction of tobacco consumption and decrease in overall lung carcinoma rates. These findings further support the hypothesis of potentially emerging, independent risk-factors of carcinogenesis, involving PRKCA upregulation. Our present data, however, do not allow a conclusion as to which factors are important.

**Figure 3:**
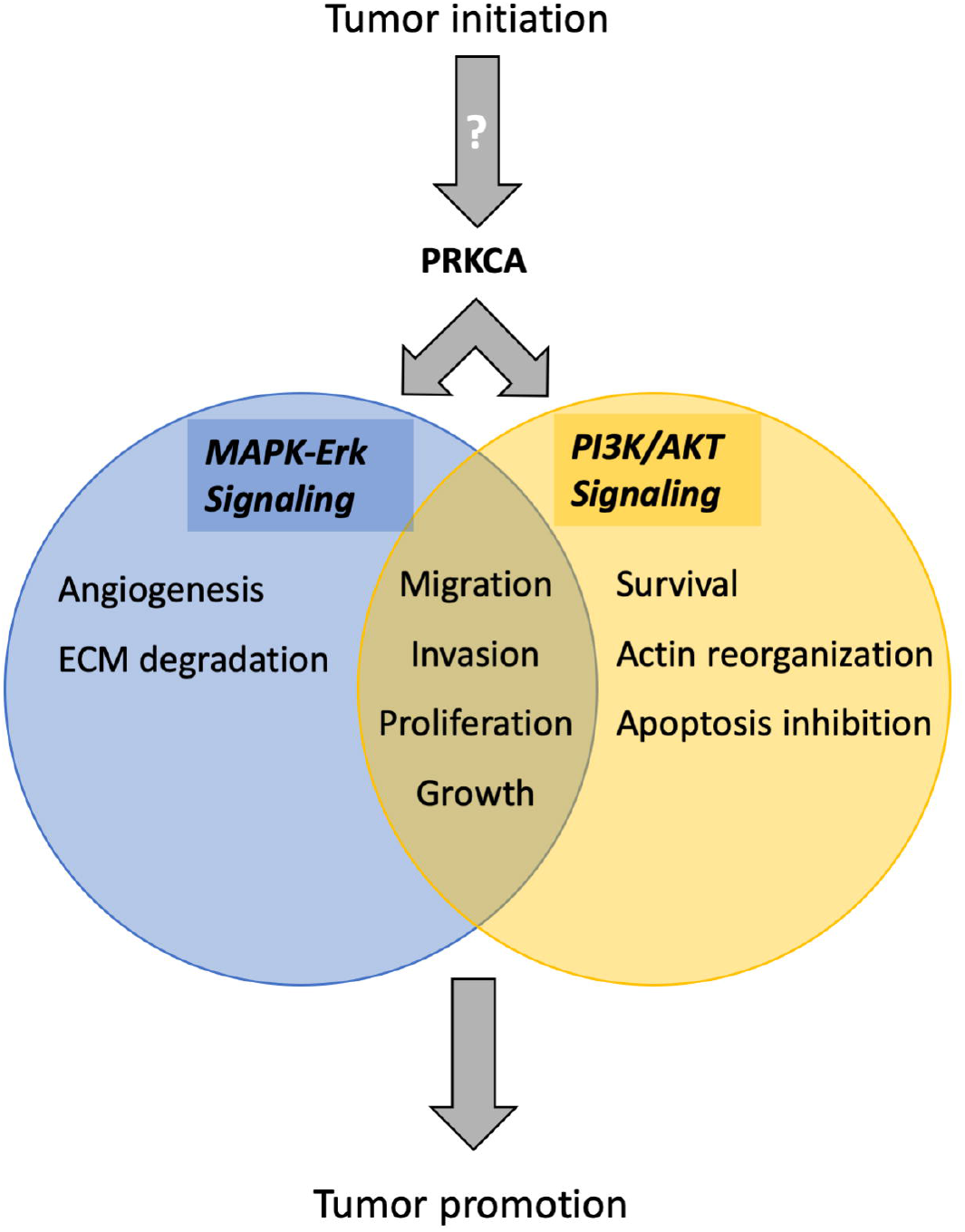
Activation of PRKCA is known to elicit downstream tumor-promoting effects via the MAPK/Erk and PI3K/Akt Signaling Pathways in several cancer types ^32–36^. Both pathways are known to promote cell growth, proliferation, migration and invasion. Additionally, the MAPK/Erk Signaling Pathway also elicits angiogenesis and degradation of the extracellular matrix (ECM), while dysregulation of the PI3K/Akt pathway can lead to actin reorganization, improved cellular survival and inhibition of apoptosis ^30,31^. The tumor-initiating mechanism of PRKCA activation and overexpression is unknown.

Our results may also have therapeutic implications since several compounds have been under clinical investigation to target overexpression of different PRKC isoforms, including PRKCA, in cancer patients. Strategies include inhibition of upstream regulators, small molecule competitive inhibitors or antisense oligonucleotides ^55^.

In conclusion, our results suggest that PRKCA overexpression may define a distinct subtype of early-onset OTSCC with poor prognosis and a yet unknown mechanism of carcinogenesis. The occurrence of distinct early-onset OTSCC subtypes and varying subtype distributions in study cohorts would offer a possible explanation for controversial reports about survival in young OTSCC patients.

## Data Availability

All relevant data regarding the Viennese study sample are included in the manuscript. Protein expression data extracted from the TCGA databank and adapted for the analysis in this study have been submitted as supplementary material. All relevant genomic and transcriptomic datasets analysed in this study are freely available for download under the public URLs provided below.

https://www.cancer.gov/tcga

https://www.cbioportal.org

## Acknowledgements

The results published here are in part based upon data generated by the TCGA Research Network: https://www.cancer.gov/tcga.

The Mouse MAB Anti-ANXA1 (EH17a) developed by Joel D. Ernst at the University of California, San Francisco, CA, US was obtained from the Developmental Studies Hybridoma Bank, created by the NICHD of the NIH and maintained at The University of Iowa, Department of Biology, Iowa City, IA 52242.

We also wish to thank Prof. Christian Schoefer and Marianne Fliesser from the Center for Anatomy and Cell Biology, Medical University of Vienna, for providing reference tissue samples for antibody evaluation.

## Abbreviations

OTSCC: oral tongue squamous cell carcinoma
TCGA: The Cancer Genome Atlas
PRKCA: protein kinase C alpha
OS: overall survival
ANXA1: Annexin 1
DFS: disease-free survival
HNSCC: head and neck squamous cell carcinoma
HPV: human papilloma virus
CNV: copy number variant
LADC: lung adeno carcinoma
AJCC: American Joint Committee on Cancer
RPPA: reverse phase protein array
WMA: World Medical Association
IHC: immunohistochemistry
EMT: epithelial-to-mesenchymal transition

